# Efficacy and Safety of Substance-Based Therapies in Anthroposophical Medicine: A Systematic Review

**DOI:** 10.1101/2025.03.26.25324709

**Authors:** Daniel Savran, Nikolas Reisecker, Fiona Tinnefeld, Justin Oosterlee, Patrick Tauber, Jan Zourek, Birgit Heller, Caroline Reitbrecht, Anita Rieder, Jutta Hübner, Edzard Ernst, Harald H. Sitte, Maria de la Cruz Gomez Pellin

**Affiliations:** Center for Physiology and Pharmacology, Institute of Pharmacology, Medical University of Vienna, Vienna, Austria; Medical University of Vienna, Department of Medicine I, Clinical Division of Haematology and Haemostaseology; University Library, Scientific Searching Service, Medical University of Vienna, Vienna, Austria; Center for Public Health, Department for Social and Preventive Medicine, Medical University of Vienna, Vienna, Austria; Klinik für Innere Medizin II, Universitätsklinikum Jena, Germany; University of Exeter, UK; AddRess Centre for Addiction Research and Science, Medical University of Vienna, Vienna, Austria; Hourani Center for Applied Scientific Research, Al-Ahliyya Amman University, 19328 Amman, Jordan

**Author notes:** Address correspondence to: Harald H. Sitte, Center for Physiology and Pharmacology, Institute of Pharmacology, Medical University of Vienna, Waehringer Strasse 13a, A-1090 Vienna, Austria (M). Department of Medicine, Naomi Berrie Diabetes Center, Irving Medical Center, Columbia University, New York, USA.

**Keywords:** Anthroposophy, Systematic reviews, Drug-Related Side Effects and Adverse Reactions, Complementary therapies, Anthroposophic Medicine, Anthroposophical Medicine

## Abstract

**Introduction:** Based on the teachings of Rudolf Steiner, anthroposophic medicine (AM) is practiced in more than 80 countries around the world today. AM blends physical, mental and spiritual dimensions of life into patient care, nevertheless regarding itself as highly scientific. As a comparable systematic review (SR) of AM conducted more than two decades ago is now outdated, this review aims to establish an updated summary of the existing clinical evidence on AM.

**Objectives:** The primary aim was to evaluate the efficacy of substance-based interventions in AM for patients with acute or chronic illnesses. Additionally, it assessed the safety and adverse drug reaction (ADR) profile of AM remedies.

**Methods:** We reviewed available evidence on substance-based AM, focusing on mortality, morbidity and safety. All randomized and non-randomized trials of AM-specific monotherapies were considered, excluding mistletoe due to existing recent high-quality reviews. A Cochrane-compliant search strategy was employed across Medline, EMBASE, Cochrane Central Register of Controlled Trials, CINAHL, PsycInfo, and Anthromedlit-Datenbank, along manual reference list searches including studies published until November 2024.

**Results:** Our searches yielded 360 hits of which 17 publications met inclusion criteria. Endpoints and diseases included allergic rhinitis, pain, stroke, and quality of life related outcomes. The only statistically significant result favoring AM was a post-hoc analysis of a neurasthenia trial—a vague diagnosis no longer included in the ICD-11. Another study assessing quality of life claimed clinically relevant improvements, but this was based on inappropriate statistical analyses. None of the other studies reported results unambiguously favoring AM. AM was well tolerated, with studies showing adverse drug reactions occurring in 0% to 3% of patients.

**Conclusion:** Twenty years after a similar systematic review, the evidence for AM treatments remains largely unchanged. We conclude that these therapies are not supported by sound evidence. Their use in routine care must therefore be questioned.

**Funding & Registration:** This SR was supported by the Medical University of Vienna, Vienna, Austria and registered in PROSPERO with the number PROSPERO 2024 CRD42024620083, available from: https://www.crd.york.ac.uk/prospero/display_record.php?ID=CRD42024620083

**Strengths and limitations of this study:**

- **Comprehensive and systematic approach**: This review adhered to PRISMA guidelines, employed a Cochrane-compliant search strategy across six databases, and included rigorous inclusion/exclusion criteria to ensure a thorough assessment of anthroposophic medicine (AM) interventions.
- **Inclusion of both randomized and non-randomized studies**: Unlike previous reviews that focused solely on RCTs, this study also considered non-randomized trials and observational studies, increasing the breadth of available evidence while acknowledging the limitations of non-randomized data.
- **Rigorous risk of bias and quality assessment**: Studies were evaluated using established tools (ROBIS, RoB 2, ROBINS-I), ensuring a transparent and critical assessment of methodological quality and potential biases.
- **Exclusion of multimodal interventions for clarity**: To isolate the effects of AM substance-based therapies, this review excluded studies that combined AM remedies with non-pharmacological interventions, allowing for clearer attribution of treatment effects.
- **Potential limitations in language and publication bias**: The review was restricted to studies in English, German, Spanish, Dutch, Czech, and Ukrainian, which may have led to the exclusion of relevant studies published in other languages or non-indexed sources. Additionally, the reliance on published literature may not fully account for negative or unpublished findings.

**Key Messages:**

- **What is already known on this topic**: AM is widely practiced and claims to integrate conventional medical practices with spiritual concepts, but its scientific foundation is highly questionable. A previous systematic review from 2004 found no high-quality evidence supporting the efficacy of AM treatments, leading to controversy between supporters of AM and critical clinicians.
- **What this study adds:** This systematic review found no robust evidence supporting the efficacy of substance-based AM interventions for any medical condition with reviewed studies suffering from significant methodological flaws, selective reporting, and inappropriate statistical analyses, undermining their credibility.
- **How this study might affect research, practice, or policy:** This calls into question their continued use in routine medical practice. Future research on AM must adhere to rigorous methodological standards to avoid misleading conclusions and ensure scientific integrity. Policymakers and healthcare providers should critically assess the inclusion of AM treatments in public healthcare systems and medical education.

## Introduction

Anthroposophic medicine (AM), an integrative system blending conventional medical practices with spiritual and mental insights, was founded by Rudolf Steiner (1861-1925) in close collaboration with the Dutch physician Ita Wegman. Steiner, an Austrian philosopher, mysticist, and theosophist, also known for his Waldorf schools and ‘biodynamic farming’, considered his spiritual philosophy a vital step in the progress of Western thought, emphasizing the interplay between the physical, mental, social, and spiritual dimensions of human life.^1–3^ Presenting his ideas as scientifically based, Steiner outlined his self-invented approaches to the concept of proof, dismissing generally acknowledged standards of evidence and claiming instead that the path leading to a discovery itself provides sufficient proof for the discovery.^4^ Through a vast number of lectures, books and articles, his work included agriculture, architecture, cosmetics, education, life-style and medicine, reflecting his vision of harmonizing the material and spiritual domains of life. Steiner and Wegman laid the foundations for AM and established clinics, research institutes, and training programs. Their text “Extending Practical Medicine” was released after Steiner’s death in 1925 and remains a cornerstone in the field of AM.^5^

Anthroposophic medicine is built on several key premises: it views health as a dynamic balance between body, soul, and spirit, influenced by the four Formative Forces of the human being, i.e., physical body, etheric life forces, astral soul, and spiritual individuality. These forces are said to be integrated into a three-fold constitution of the human organism, consisting of three major systems: the “nerve-sense system”, the motor-metabolic system, and the rhythmic system, each predominating in their own regions of the body and providing the conditions for various physiological and emotional functions.^1,6^ This mereology of body constitution is incommensurably at odds with our current understanding of present-day scientific medicine, a fact that is acknowledged even by AM-advocates.^7^

The anthroposophic diagnostic approaches integrate conventional methods with anthroposophical insights, including detailed patient narratives, observations of gestures and physiognomy, and an emphasis on individual constitution.^8^ Anthroposophic therapies include medications prepared from plants, minerals and organic specimen through unique pharmaceutical processes, artistic therapies like painting and music, and eurythmy, a movement therapy.^9^ They aim to stimulate the patient’s self-healing capacities and are often employed alongside conventional or other alternative interventions.^10^ In this context, Steiner often speaks of homeopathy, albeit not exactly the method put forward by Samuel Hahnemann. Steiner employs homeopathy merely as the rhythmic breakdown of a substance, which he termed “homeopathization”, which he claimed would unlock its medicinal healing powers.^11^

A 2004 systematic review^3^ assessed all randomized clinical trials (RCTs) evaluating the efficacy of AM, both as singular and add-on to science-based medical practices. Since none of the 112 identified studies met the stringent inclusion criteria of randomization, the author concluded that “the question of whether the anthroposophical approach does more good than harm cannot be answered”. A letter to the editors issued by prominent voices of the anthroposophical community then criticized this review claiming that it contains “many misleading and false statements”, and calling it “unacceptable by scientific standards”. ^12^ However, Steiner’s explicit rejection of experimentation as the foundation of knowledge might explain that randomized studies of high quality were not found in the SR by Ernst.^3,13^ Anthroposophy’s history is also not without controversy. Steiner’s writings contain passages that show that racist and anti-Semitic concepts had an important place in the worldview of Anthroposophy, which he developed.^14–17^ Under National Socialist rule, Anthroposophy as an organized movement was suppressed and some of its followers were persecuted, but there were also prominent Nazis who supported elements of its teachings, and many followers managed, to varying degrees, to arrange themselves with the regime.^16,17^ This historical and ideological complexity continues to shape the perception and practice of AM, which today is practiced in around 80 countries worldwide^18^. Understanding its roots, principles, and controversies is essential for an informed evaluation of its place in contemporary healthcare.

The present systematic review is aimed at overcoming any flaws of previous assessments and at providing a rigorous summary of the evidence for or against AM.

## Methods

### Objectives

The primary objective of this systematic review was to determine whether substance-based therapies in AM are, according to standardized and validated measures, effective in patients with acute or chronic illnesses. A further objective was to assess the safety and adverse drug reaction (ADR) profile of AM remedies.

This systematic review was written in accordance with the PRISMA checklist.^19^

### Types of Interventions

Identifying, measuring and interpreting treatment effects in populations receiving multiple interventions poses a great challenge. To mitigate confounding factors of pharmacological and non-pharmacological approaches on the validity of our analysis and conclusions, we decided to focus exclusively on the effect of AM remedies on clinical outcomes. We considered all AM-specific substance-based treatments excluding mistletoe therapy, which has recently been extensively reviewed by Staupe et al.^20^, Hofinger et al.^21^, Ostermann et al.^22^ two SRs by Loef et al.^23,24^ and further tested in a RCT by Wode et al^25^.

### Criteria for Study Designs

A comprehensive list of all inclusion and exclusion criteria is provided in the supplementary information. For the evaluation of efficacy, we included RCTs and non-randomized clinical trials as well as systematic reviews. Interventions used in the trials had to be clearly defined and limited to one AM-preparation as a mono-therapy. Because multimodal interventions (e.g. multiple AM-preparations or non-pharmacological interventions, such as eurythmy, art therapy etc.) do not allow a clear attribution of cause and effect, such studies were excluded. If studies used multiple interventions in multiple different cohorts, results had to be reported separately for each cohort with a distinct intervention, otherwise the study was excluded. The control groups had to either receive placebos or active medications and studies with a no treatment control groups were excluded.

Outcome measures had to have unambiguous clinical relevance, e.g. mortality, morbidity, quality of life or activities of daily living. Studies reporting surrogate parameters only (e.g. laboratory parameters, changes in ECGs) were excluded. No limitations were set in terms of duration of intervention, follow up times, age, participant characteristics, and setting (i.e. inpatient or outpatient). Studies within the native linguistic scope of the reviewers were included (i.e. English, German, Spanish, Dutch, Czech, Ukrainian).

Regarding our assessment of safety, we additionally included observational prospective and retrospective studies with or without control groups.

### Literature Search Strategy

A table with search terms corresponding to the individual sub-items in the PICOS scheme was created for the search, in accordance with the methodology as defined in the Cochrane Handbook for Systematic Reviews of Interventions ^26^. The information specialists of the systematic review consultation service of the Library of the Medical University of Vienna assisted in designing database specific search strings. The following databases were searched: Medline, EMBASE, Cochrane Central Register of Controlled Trials, CINAHL, PsycInfo, and Anthromedlit-Datenbank. Moreover, we reviewed the reference lists of the included studies and of systematic reviews for relevant publications. Unpublished studies were not included as well as conference proceedings, trials registers, internet resources and contact with investigators, experts and manufacturers. Tables with the full database specific search strings are provided in the supplementary information. We included studies published until November 2024.

### Data collection and extraction

Study selection was conducted in a two-step process. Firstly, two reviewers independently screened titles and abstracts of identified papers using the online platform Rayyan^©^ ^27^. In case of agreement, the study either automatically entered the second stage or was excluded. In case of disagreement, an external third reviewer was consulted to bring about a consensus. Secondly, inclusion/exclusion was determined based on the full text of each paper that passed the first hurdle and handled analogously to the first step. Both stages were conducted in a blinded manner and unblinding was done after each reviewer had submitted all votes. From the second step on, every exclusion was documented with supporting arguments, which can be found in the supplementary information.

The number of identified, selected and excluded publications at each step is visualized in a PRISMA diagram (Figure 1).

**Figure 1:**
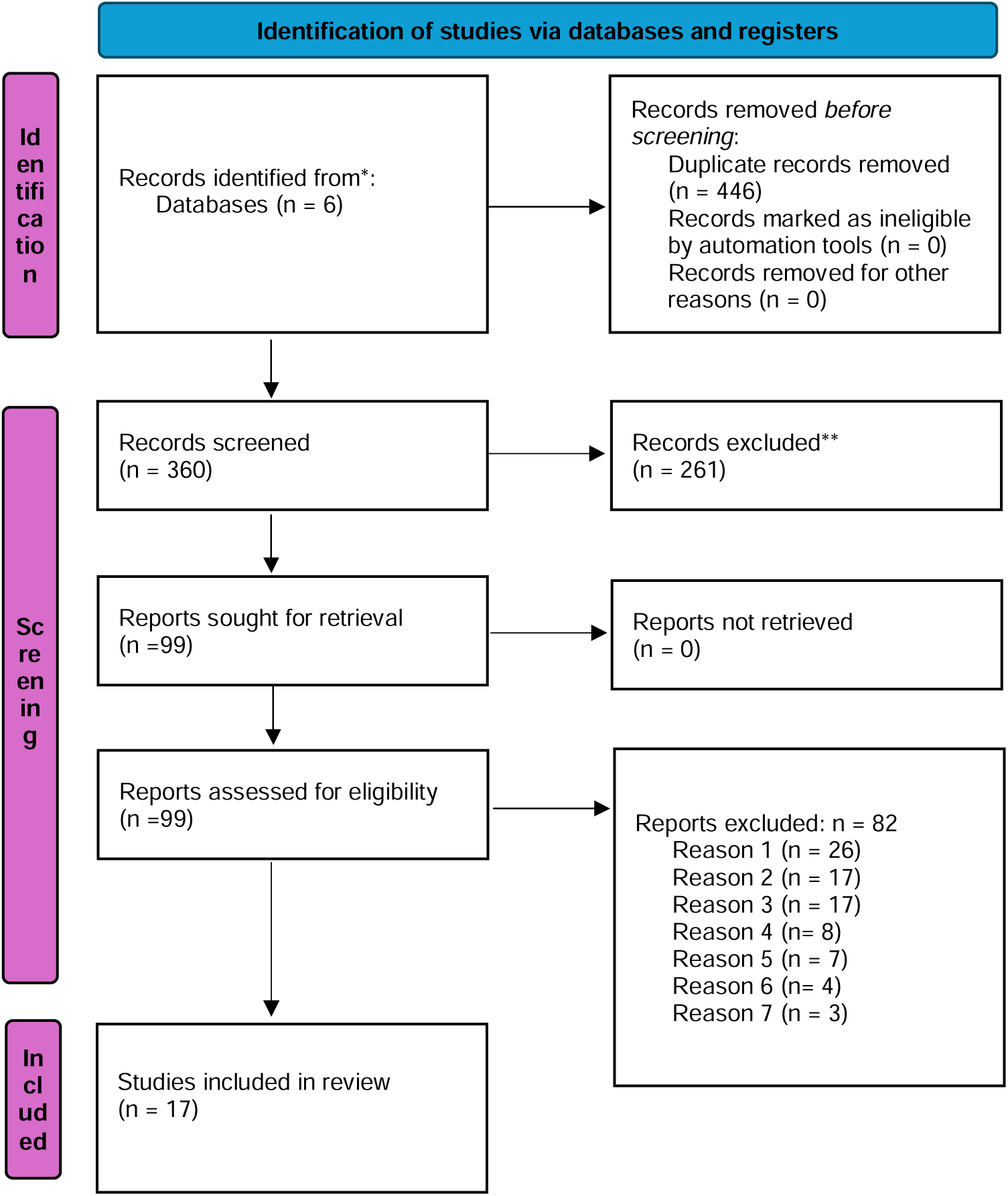
PRISMA 2020 flow diagram for new systematic reviews which included searches of databases and registers only. Reason 1 = wrong publication type Reason 2 = missing pharmacological/clinical intervention Reason 3 = multimodal intervention Reason 4 = no control/placebo group Reason 5 = wrong outcome measure Reason 6 = mistletoe Reason 7 = not AM specific Source: Page MJ, et al. BMJ 2021;372:n71. doi: 10.1136/bmj.n71. This work is licensed under CC BY 4.0.

All included studies were subjected to quality assessments. Relevant key data were extracted into predefined standardized tables independently by two reviewers. These included: Title, authors, year of publication, journal, study design, control group, population, outcomes and their measurements, results and author’s conclusions. The resulting tables were assessed for completeness and accuracy and then combined into one. For major discrepancies, a third reviewer was consulted to generate a consensus.

### Risk of Bias and Quality Assessment

Depending on the three major study designs, the following tools were used to assess the methodological quality and risk of bias of all included papers:

– Systematic reviews and meta-analyses were evaluated with the ROBIS tool (Risk of Bias in Systematic Reviews) ^28^
– RCTs were analysed using the Cochrane RoB 2 tool (Risk of Bias 2) ^29^
– Non-randomized studies were assessed with the ROBINS-I tool (Risk Of Bias In Non-randomised Studies - of Interventions) ^30^

Analogous to previous steps, assessments were done blinded by two independent reviewers, compared and discrepancies were resolved by a third reviewer.

### Strategy for data analysis

In case of identification of several comparable studies, a meta-analysis was planned, including a heterogeneity test to enable a valid comparison of results. Alternatively, a representation as I^2^ according to Higgins and Thompson ^31^ or Q according to Cochran ^32^ was intended. However, due to the low abundance of included studies and overlapping endpoints/interventions no such analyses were performed.

### Patients and Public Involvement

No patients or the public were involved in designing, analyzing and writing of this review.

## Results

After removal of duplicates, the initial searches resulted in a total of 360 abstracts. After screening, 113 abstracts were identified for full-text scrutiny. A total of 17 papers passed this hurdle and thus could be included in the final analysis. Five RCTs ^33–37^, two non-randomized trials^38,39^ (Table 1), five prospective pharmacovigilance studies^40–44^, one retrospective pharmacovigilance study^45^ (Table 2) and four systematic reviews ^6,46–48^ were finally used for data extraction. Due to the profound heterogeneity of the reported interventions and treated conditions, an insightful meta-analysis was not feasible.

**Table 1:** Overview and summary of intervention studies and their most relevant results. Abbreviations: ITT – intention to treat; PP – per protocol; LQNS - Lemon and quince nasal spray; MRS II - Menopause Rating Scale II; KPC - Kalium phosphoricum comp.; PSQ – Perceived Stress Questionnaire; WOMAC - Western Ontario and Mc. Masters University Arthritis Index; ETG – experimental transdermal gel; PTG – placebo transdermal gel;

| Name | Year | Study Type | Population | Disease/<br>Symptoms | Outcome | Intervention | Control/Comparator | Result |
| --- | --- | --- | --- | --- | --- | --- | --- | --- |
| Hoffmann A. et al.<sup>33</sup> | 2016 | Randomized, placebo controlled, cross-over study | Total PP: n = 31<br>Total ITT: n = 41 | hay fever symptoms | **Primary Outcome:** inspiratory nasal flow at 150 pascal measured with rhinomanometry 10 and 20 min after provocation with grass pollen after 1 week of treatment | **Lemon and quince nasal spray** (Gencydo® from Weleda) containing: 80-120 mg/10mL lemon juice, 300 mg/10mL quince extract, saline 0.9% Administered 3x/day with two sprays (2x140 mcg) in each nostril for seven days and one spray 1 hour prior to control visit. | Placebo: identical appearance & scent - different taste - | **ITT analysis:** No significant difference in nasal flow between LQNS and placebo.<br>**PP analysis:** LQNS improved pre-provocation inspiratory nasal flow after one week (p=0.032).<br>Post-provocation: lower relative reduction in inspiratory flow (-7.78% ± 24.02p=0.059) and lower relative increase of inspiratory resistance (71.43% ± 198.49p=0.044). |
| Von Hagens et al.<sup>34</sup> | 2012 | randomized, placebo controlled, double bind, three armed trial | Group 1 n = 30<br>Group 2 n = 32<br>Group 3 n = 32<br>no washout phases | menstrual symptoms | **Primary outcome:** changes of the MRS II 12 after 12 weeks by pooling groups 1+3 and comparing to group 2 (placebo) | 3x10 globuli daily of **Apis regina tota GL D4, Argentum metallicum D5, Ovaria bovis GL D4** | placebo | **MRS II** reduction after 12 weeks: (p=0.441)<br>**Intervention:**-1.4 (SD 5.6, 95% CI - 2.8 to 0), Group 1+3 n = 62<br>**Placebo:** -2.3 (SD 5.8, 95% CI -4.4 to -0.3), Group 2 n = 32 |
| Hellhammer J et al.<sup>35</sup> | 2024 | randomized, double-blind, placebo-controlled parallel group phase IV | KPC group: n = 77<br>Placebo group: n = 77 | neurasthenia", meaning exhaustion of the "nerve strength" | **Primary outcome:** Neurasthenia symptoms after 6 weeks, assessed by nervous exhaustion score (0–3 scale, 12 symptoms), PSQ, and SF-36 Health Survey. | **Kalium phosphoricum comp.** (Manufacturer: Weleda AG, Schwäbisch Gmünd, Germany) | placebo | **Mean symptom sum score** did not differ significantly between KPC and placebo (10.6 ± 5.7 vs 10.9 ± 6.0, p=0.728) **mean PSQ** (KPC: 41.9 ± 18.0, placebo: 40.0 ± 18.3, p=0.505)<br>**SF-36** showed no significantly stronger improvement at follow up, neither of the physical nor the mental sum scores for the KPC arm when compared to placebo(physical: KPC: 46.9 ± 9.0, placebo: 47.3 ± 9.0, p=0.773; mental: KPC: 43.1 ± 12.0, placebo: 44.4 ± 12.0, p = 0.510) |
| Huber R. et al.<sup>37</sup> | 2010 | Prospective, randomized, double blind, placebo controlled study | AG5: n= 45<br>Placebo: n= 44 | osteoarthritis of the knee - pain | **Primary outcome:** WOMAC pain score after 4 Weeks of treatment | **Articulatio Genus D5**, s.c. injection 3x per week in affected knee at two trigger points (points painful at palpation) if available, otherwise above the medial and lateral ligaments of the knee 0.5ml per trigger point | Placebo: solvent of AG5, an isotonic solution of sodium hydrogen carbonate and sodium chloride | From baseline to day 28 **WOMAC pain scores** improved significantly *within* both groups (each $p \leq 0.001$)<br>Baseline vs follow up:<br>- **WOMAC pain score AG5:** $5.1 \pm 1.8$ vs $3.2 \pm 2.2$<br>- **WOMAC pain score placebo:** $4.3 \pm 1.5$ vs $3.0 \pm 1.8$ However, AG5 showed no significant superiority over placebo in reducing pain scores ($\Delta=0.33$, 95% CI: $-11.8$ to $5.3$, $p = 0.459$) |
| Ghelman R. et al.<sup>36</sup> | 2020 | randomized, double-blind, placebo-controlled, phase 2 prospective clinical trial | ETG : n=10<br>PTG n=10 | post poliomyelitis syndrome | **Primary outcome:** Pain intensity (VAS), multidimensional pain evaluation (McGill Pain Questionnaire), and thermographic assessment (FLIR T420 infrared camera). | nightly application of the **ETG** in groups A and C, at a dose of 1 g in the painful regions for 12 weeks. The **ETG** vials had a final concentration of 10% of the active ingredient | nightly application of PTG in groups B and D, at a dose of 1 g in the painful regions for 12 weeks. PTG vials contained only the inert excipient soy lecithin. | **VAS pain reduction:** ETG: 20.7% vs. placebo: 24.8% (not significant).<br>**McGill Pain Questionnaire:** Not reported. **Thermography:** Significant thermal asymmetry; affected limb hypothermia; hyperthermal signals in osteoarthritis/sacroiliitis. Systemic findings suggestive of sleep disturbances, gastrointestinal dysfunction, and stress-induced thermogenesis. |
| Wilkins et al.<sup>38</sup> | 2008 | prospective non randomized cohort trial | Naja comp.: n = 82<br>Control: n = 123 | stroke | **Primary outcome:** changes in the Barthel Index between admission and discharge | additionally to usual treatment - injections of **Naja comp** 2-3x/week (Crotalus terr. D20, Lachesis D12, Naja D10, Vipera berus D30) | usual evidence based treatment | **Barthel Index Baseline** (interventions vs control): $31,3 \pm 28,7$ vs. $53,9 \pm 32,5$ Discharge: $52,7 \pm 33,9$ vs $72,1 \pm 31,2$ **Total change:** $21,4 \pm 19,6$ vs $17,9 \pm 17,9$ no in-between group analysis/statistic was reported |

**Table 2:** Overview and summary of ADR studies. Abbreviations: ADR – adverse drug reaction; AM – anthroposophic medicine; CAM – complementary and alternative medicine, CON – conventional medicine; MDADs - maximum daily administration doses

| Name | Year | Study Type | Population | Outcome | Intervention | Result | Author's Conclusions |
| --- | --- | --- | --- | --- | --- | --- | --- |
| Jeschke E. et al.<sup>42</sup> | 2009 | Prospective, multicenter observational study. | **Patients:** n = 50115<br>**total prescriptions:** n = 199 387 of these, 18 830 (37.6%) received 25 652 prescriptions (12.9%) with a total of 42 378 remedies (11.8%) containing Asteraceae extracts | **ADRs:** Classified by severity (serious/non-serious, ICH criteria), causality (Uppsala Monitoring Centre), system organ class, and frequency categories | Any product containing Asteraceae | **No serious ADRs reported; 11 non-serious ADRs** (0.16% of 6961), linked to homeopathic dilutions (10xD1/1xD4).<br>ADRs were linked to products with Echinacea spp., Calendula officinalis, Matricaria recutita, Artemisia abrotanum, Arnica montana, Silybum marianum, Taraxacum officinale and Solidago virgaurea<br>**Relative Risk of ADR** while using Asteraceae 0.13 (95% CI 0.07, 0.23)<br>**PRR for Asteraceae** with respect to all other prescriptions:<br>**PRR for skin disorders** (7 ADRs): 1.69 (95% CI 0.97–1.96); **for GI disorders** (3 ADRs): 0.98 (95% CI 0.34–3.61) (3 ADRs) none of PRR results were significant according to criteria developed by Evans et al. | This study provides a systematic overview of prescribing patterns and ADRs for Asteraceae-containing remedies in the German primary-care sector. Asteraceae-containing remedies were used frequently in this context, especially among children. Most of the ADRs observed in the present study involved skin disorders and subcutaneous tissue complaints. Our results indicate that treatment with Asteraceae-containing remedies is not associated with a high risk of ADRs. The data in this regard are especially clear for Arnica montana, Calendula officinalis, Matricaria recutita and Taraxacum officinale. For the other Asteraceae (e.g. Silybum marianum, Artemisia abrotanum and especially Solidago virgaurea), a larger sample with more prescriptions is needed to assess ADR risk properly. |
| Tabali et al.<sup>40</sup> | 2012 | ADRs: prospective, multicenter, observational study | EvaMed Pharmacovigilance Network:<br>total patients treated: 88.431 (f: 49.964, 56,5%) total treatments prescribed: 1.018.626 | any ADRs during observation period from January 2004 to June 2009<br>**ADR data:** seriousness, severity, system organ classes, causality, management, outcome | any AM remedy treatment | **total ADRs** n = 430; 18 were classified as off label use<br>**final count:** 412 ADRs in 389 patients<br>**total CAM associated ADRs** = 124 (30,1%) total severity: Grade I = 15,0%; Grade II = 75,0%; Grade III = 9,2%; Grade IV = 0,7%; | In a small sample of 7 CAM physicians who reported both serious and nonserious ADRs, there were 213,900 CAM prescriptions (54.5%) of 392,243 total prescriptions for CAM and CON drugs. The serious ADR rate was 0.6 per 10,000 prescriptions for CON drugs, and no CAM ADRs were serious. The overall ADR rates per 10,000 prescriptions were 4.4 for CAM prescriptions and 13.0 for CON prescriptions. |
| Hamre, H.J. et al.<sup>41</sup> | 2017 | prospective pharmacovigilance study | n=44,662<br>**Prescriptions:** 717,545<br>**total AM remedies prescribed** = 311,731 (1722 different remedies) | **Primary:** the frequency of ADRs to AM remedies [any ADR, ADRs Grade III–IV and serious ADRs (SADRs)], relative to the number of AM remedy prescriptions. | any AM remedy treatment | **ADRs** of any intensity occurred in 0.071% of AM remedy prescriptions and in 0.502% of patients prescribed AM remedies. **ADRs with intensity Grades III–IV** occurred in 0.002% of AM remedy prescriptions and in 0.016% of patients prescribed AM remedies. | This analysis of more than 40,000 outpatients in a prospective cohort study confirms previous findings from other studies with\ 1000 patients. ADRs to AM remedies were rare (0.071% of AM remedy prescriptions), ADRs of high intensity and SADRs were very rare (0.002 and 0.0003%, respectively). |
| Jong MC. et al.<sup>45</sup> | 2021 | Retrospective Cohort Study | 4 Manufacturers of Anthroposophic medicinal products. number of Patients: 2765 | **ADR analysis included:** Report date, report type, reporter qualification, ADR description (MedDRA), seriousness (MAH classification), labeling status at reporting, outcome (per reporter), and causality (MAH) | any AM remedy treatment | **ADR frequency:** 1.5 ADRs per million MDADs; **serious ADRs:** 0.03 per million MDADs. **Causality:** Most classified as “possible,” with resolution in most cases; **no fatal ADRs** reported. Reporting: >99% spontaneous; 5,506 ADRs in 2,765 patients, more frequent in women and older adults. **Affected products:** 370 anthroposophic medicinal products. **Common ADRs:** Local/parenteral AM remedy-application-related; oral AM remedies—nausea, rash, pruritus. **Most affected organ classes:** Administration site, eye, skin/subcutaneous tissue. **Serious cases:** 5 reports of possible herb-induced liver injury. | Retrolective analysis of pharmacovigilance data from spontaneous reporting systems is an important method to investigate and monitor the safety of AM therapy in everyday practice. In the present study, ADRs related to AM therapy were very rare and, as in previously published studies, mostly reversible and non-serious in nature. This applied to the whole dataset and all subgroups analysed. Notably, the frequency of ADRs in this study based on spontaneous reporting (1.5 ADR per million MDADs) is not directly comparable to frequencies in prospective clinical studies nor to frequencies based on other measures of patient exposure than MDADs, such as numbers of users or prescriptions. |
| Hamre, H.J. et al.<sup>39</sup> | 2007 | prospective observational comparative study | **Patients prescribed AM remedies for chief complaint** (Day 0-28): 715/715<br>**Patients prescribed non-AM remedies for chief complaint** (Day 0-28): 377/715 | **Follow-up:** Days 7, 14, and 28; final interview at 16.2±8.5 days. Patients with full recovery by Days 7 or 14 exited the study. **ADR assessment:** Classified by seriousness, severity, system organ class, causality, management, and outcome. | any medication produced by the pharmaceutical companies Weleda AG, Arlesheim, Switzerland or Wala-Heilmittel GmbH, Eckwälden, Germany. | **AM Medications:** 327; **Total applications** = 73.443 **Confirmed ADR** to anthroposophic medicines in 2 Patients (0.28%) (bronchopneumonia, acute tonsillitis), 2 Medications (0.61%) (Pneumodoron caused sleeplessness, Prunus spinosa - mild swelling and redness at the injection site), 3 Applications associated with ADR (0.004%), ADR in 0.3% of patients and 0.004% of applications; **Median duration** 2 days | In this prospective study of 715 outpatients with acute respiratory and ear infections, we found a low frequency of ADR and no serious ADR to AM remedies. Study results suggest that short-term AM therapy for acute respiratory and ear infections is well tolerated. |
| Hamre H.J. et al.<sup>43</sup> | 2006 | Prospective observational cohort study | 662 outpatients (ages 1–75) with chronic diseases; 949 different anthroposophic medications used over 11,487 patient-months. | Number of patient-months for all medication within 2 years after enrollment. **ADR data:** frequency, diagnosis, intensity, seriousness, causal relationship to medication, most probable cause, actions taken against ADR, outcome of ADR | Any anthroposophic medication prescribed in 131 medical practices across Germany. | **ADRs documented:** 1,861, mostly non-specific symptoms (27.6%), musculoskeletal (16.9%), respiratory (8.2%), and digestive diseases (6.6%). **No serious ADRs** related to AM remedies occurred. Among 284 suspected ADRs, **55 were linked to AM remedies** in 29 patients. **Confirmed ADRs** affected 3.0% of AM remedy users, with an incidence of 1 ADR per 382 patient-months. Two patients (0.3%) experienced severe ADRs, but none were serious. Low overall frequency of ADRs, with most being mild to moderate in severity. | In this 2-year prospective study of 662 outpatients with chronic disease, we found a low frequency of ADRs and no serious ADRs to AM remedies. Study results suggest that long-term AM therapy is generally well tolerated. |
| Süsskind M. et al.<sup>44</sup> | 2012 | Prospective observational study | 3,813 hospitalized patients at Community Hospital Havelhöhe, Berlin, Germany, over six months (May–November 2007). | **Type A** (dose dependent and predictable) and **Type B** (dose independent and unpredictable) ADRs according to Edwards and Aronson, **ADR data:** start, duration and outcome, suspected drug(s), and management strategy, organ manifestation (WHO | Any medication during the hospital stay | **ADRs documented** in 174 patients (4.6%) which experienced 211 ADRs, with 57 (27.0%) classified as serious. **Six ADRs** (2.8% of total ADRs) were linked to AM remedies affecting 0.27% of AM remedy-exposed patients. These included mild (CTCAE grade 2) local skin reactions and fever from mistletoe injections, and one maculopapular exanthema case after *Fragaria vesca/Vitis vinifera*. **No serious ADRs were reported.** | Despite the skepticism towards CAM and AM remedies, our data show that ADR surveillance is possible and a reasonable safety profile of AM remedies can be assumed. ADRs associated with allopathic medication detected in our study were comparable with regard to incidence and nature to published data, underlining the comprehensiveness of our prospective survey. In summary, our results suggest that ADRs caused by AM remedies occur only with a low incidence and produce mainly mild to moderate symptoms. Since mistletoe extracts were the predominantly |
|  |  |  |  | SOC-code), causality,<br>severity, seriousness |  |  | used AM remedies in this cohort of patients, most ADRs were associated with mistletoe extracts. This study showed the difficulties to comprehensively document the complete medications of all patients. Thus interpretation of our results must be done with great caution. |

The following four subchapters each summarize one of the endpoint categories identified: pain; quality of life/perceived well-being; improvements in stroke rehabilitation; and adverse drug reactions/safety.

### Pain

In a four-arm, RCT, Ghelman et al. evaluated the efficacy and safety of an AM for chronic pain in post-polio syndrome (PPS) outpatients. ^36^ Forty-eight patients were randomized into four groups receiving either a transdermal gel (TG, *containing Rhus toxicodendrum D4 (1.66%), Arnica montana D3 (1.66%), Apis mellifica/ Atropa belladonna D3 (0.83%/0.83%), Mandragora officinarum D3 (1.66%), Aconitum napellus D4 (1.66%), and Hypericum perforatum D3 (1.66%*), a placebo gel (PTG), or a combination of either TG or PTG with weekly non-pharmacological anthroposophical clinical interventions. For the present analysis, only results reported separately for the TG and PTG arm are considered. Pain reduction was assessed using a visual analogue scale (VAS), the McGill questionnaire, and, additionally, thermography was performed. The study found no statistically significant reduction in the amount of pain in the groups with TG and PTG. The authors report no treatment-related adverse events.

Huber et al. conducted an RCT^1^ evaluating the efficacy and safety of subcutaneous injections of an AM cartilage preparation (*Articulatio Genus D5, AG5*) for knee osteoarthritis. ^37^ In total, 91 patients were randomized, with 46 receiving AG5 and 45 receiving placebo injections over four weeks, three injections per week. The primary outcome, assessed using the WOMAC pain score, showed significant pain reduction in both groups (mean from 5.1+/−1.8 to 3.2+/−2.2 with AG5 and from 4.3+/−1.5 to 3.0+/−1.8 with placebo; p < 0.001); however, AG5 did not demonstrate superiority over placebo regarding pain relief. There was, however, a significant reduction of pain medication usage in the intervention group. No severe ADR were reported, local irritation at the site of injection occurred in one patient of either group, and tolerability was rated as good or excellent by 98% of participants.

### Quality of Life/Perceived Well-Being

Two RCTs evaluated the efficacy of AM on quality of life-related outcomes. ^34,35^. In a three-period crossover trial, Von Hagens et al. treated three groups of patients (n=30; 32; 32) experiencing menopausal symptoms with either AM (*Apis regina tota D4, Argentum metallicum D5, and Ovaria bovis D4*) for two weeks, or placebo for one week. After 12 and 24 weeks, no significant differences for the primary endpoint – changes in Menopausal Rating Scale II (MRS II) scores – were observed. Clinically relevant improvements were claimed after 36 weeks in groups receiving AM for two consecutive periods, when the change during the placebo phase was included, but this finding was not supported by statistical analysis.

Hellhammer et al.^35^ investigated the effects of Neurodoron (*Kalium phosporicum comp (KPC)*) – on symptoms of Neurasthenia, a diagnosis that was abandoned in the ICD-11, describing chronic fatigue without organic cause.^49^ None of the multiple primary endpoints – i) symptom sum scores of twelve characteristic symptoms, ii) Perceived Stress Questionaire (PSQ) and iii) Short Form-36 Health Survey (SF-36) - differed significantly between KPC (n=77) and placebo (n=77) after 6 weeks. However, a post-hoc analysis showed significant improvements in the sub-scores for irritability and nervousness (characteristic symptoms), as well as joy (PSQ) in the KPC group, while no differences were found for SF-36 sub-scores.

### Stroke

One non-randomized, two-center cohort study ^38^ investigated the effects of Naja comp. injections (*Crotalus terrr. D20, Lachesis D12, Naja D10, Vipera berus D30*), a preparation based on 4 different snake venoms, on improvements of autonomy in stroke rehabilitation patients. Only patients from center II were considered for this review, since center I patients received additional homeopathic treatment. Compared to the control group (n=123) receiving conventional treatments, the addition of Naja comp. (n=82) did not result in statistically significantly higher improvements, as quantified by the Barthel-Index.

### Allergic Rhinitis

In a crossover RCT Hoffmann et al. tested the effect of Gencydo® (*C.limon succus, quince extract and saline; n=22*) vs placebo (n=21) on inspiratory and expiratory nasal flow, resistance, and symptom score after provocation with grass pollen.^33^ Nasal flow parameters and scores were acquired at baseline and after seven days of treatment, with a seven day washout period before crossover. The intention to treat analysis showed no significant differences between the groups. However, in the per protocol analysis participants treated with Gencydo® had higher absolute inspiratory flow after provocation, higher pre-provocation inspiratory flow rates and a lower relative increase of inspiratory resistance ten minutes after provocation. Absolute expiratory flow was significantly higher after 20 minutes in the intervention group. All other parameters showed no significant changes. The authors stressed, that a total of ten patients had been excluded in the per protocol analysis due to upper respiratory tract infections or major protocol violations. Nevertheless, they attribute an antiallergic effect of Gencydo®.

### Adverse Drug Reactions and Safety

Hamre et al. conducted a two-year prospective cohort study on the safety of anthroposophic medications in 662 outpatients across 131 German medical practices.^43^ Patients used 949 different AM remedies over 11,487 patient-months. A total of 1,861 ADRs were recorded, mainly non-specific symptoms (27.6%), musculoskeletal complaints (16.9%), respiratory issues (8.2%), and digestive disorders (6.6%). Of these, 284 ADRs (15.3%) were suspected ADRs, with 55 linked to AM remedies in 29 patients. Confirmed ADRs were found in 20 patients, involving 21 AM remedies. Reactions included local application site issues (6 patients), systemic hypersensitivity (1 patient), and exacerbation of pre-existing conditions (13 patients). Two patients experienced severe ADRs, but none were classified as serious. The median ADR duration was 7 days (range: 1–39). The overall ADR rate was 2.2% of AM remedies used, affecting 3.0% of users, with one ADR per 382 patient-months of AM remedy use. The authors concluded that AM therapy was generally well tolerated, with a low ADR frequency and no serious ADRs reported.

Another prospective observational study by Hamre et al. evaluated ADRs in patients receiving AM for acute respiratory and ear infections^39^. A total of 715 patients were prescribed AM, while 377 received non-AM treatments. Adverse events were assessed by interviews at Days 0, 7, 14, and 28, with follow-up ending upon full recovery. There was no in person evaluation of the reported ADRs, all data were based on patient’s subjective responses via telephone. Among 73,443 AM applications of 327 different AM preparations, ADRs were noted in two patients (0.28%) and linked to two different AM medications (0.61%): Pneumodoron (ADR=sleeplessness) and Prunus spinosa (ADR=swelling and redness of injection site). In total three applications (0.004%) were linked to ADRs, with two applications (0.003%) causing an ADR of severe intensity. The median duration of the ADRs was two days. The authors concluded that short-term AM therapy for acute respiratory and ear infections is well tolerated.

Süßkind et al. conducted a six-month prospective observational study at the Community Hospital Havelhöhe in Berlin, Germany, to assess adverse drug reactions associated with anthroposophic medications.^44^ Among 3,813 hospitalized patients, 174 (4.6%) experienced 211 ADRs, with 57 (27.0%) classified as serious. The median age of affected patients was 72 years, and 62.1% were female. Only six ADRs (2.8% of all ADRs) were linked to AM remedies, affecting 0.27% of AM remedy-exposed patients. These reactions were mild (CTCAE grade 2) and included three local skin reactions with two patients also developing fever following mistletoe injections, one patient experiencing a local skin reaction following subcutaneous Equisetum/Formica injection, one pectoral drug eruption five hours after oral Gentiana, Bryophyllum, and metamizole administration, as well as one case of maculopapular exanthema after oral administration of Fragaria vesca/Vitis vinifera. No serious ADRs were reported. The authors concluded that ADRs associated with AM remedies were rare and limited to mild symptoms, supporting a favorable safety profile for anthroposophic treatments in hospitalized patients.

Jeschke et al. conducted a prospective observational study to assess the safety profile of Asteraceae-containing remedies, a plant group commonly used in AM and herbal medicine.^42^ ADRs were documented by physicians from the German Association of Anthroposophical Physicians (GAÄD) in the EvaMed Pharmacovigilance Network from 2004 to 2006. While a small subgroup (7/38) reported all suspected ADRs, most physicians (31/38) documented only serious ADRs. Among the 18,830 identified patients who received Asteraceae-based remedies, non-serious ADRs were reported for a subgroup of 6961 patients. Only 11 (non-serious) ARDs were reported. Reactions affected the skin and subcutaneous tissue (7/11), the gastrointestinal system (3/11), and the eye (1/11). Causality was deemed probable in 6 cases and possible in 5. The estimated relative risk for Asteraceae-containing products was 0.13 (95% CI: 0.07–0.23), though proportional reporting ratio analysis yielded no significant results.

A subsequent study ^40^ with a larger sample size (n = 88,431) used the same database but analyzed a time span of over 5 years (2004-2009). The investigators found that all serious ADRs (n = 14) were linked to conventional drugs and none to AM. Overall, 124 non-serious ADRs were observed in relation to AM prescriptions. The most frequently affected organ systems were the skin and subcutaneous tissue followed by gastrointestinal tract. In their interpretation, the authors highlight the relative safety of AM medicines in comparison to conventional remedies.

Another prospective observational study ^41^ with a large sample size (n = 44,662) found that ADRs following treatment with AM were rare. ADRs during the observation period (January 2001-December 2011) were recorded in the same Pharmacovigilance Network as above. Only 0.071% of prescriptions resulted in ADRs, and a mere 0.002% led to severe ADRs. In total, 100 cases of ADRs were reported to be certainly, probably or possibly associated to AM, affecting 95 out of 44,662 patients. The most frequently reported symptoms affected the skin and subcutaneous tissue (21.6% of symptoms, n = 30/139), psychiatric conditions (19.4%), the gastrointestinal tract (17.3%), the administration site (9.4%), and the respiratory tract, including thoracic and mediastinal complaints (6.5%).

Jong et al. retrospectively analyzed pharmacovigilance data from four German distributors of AM preparations from 2010 to 2017.^45^ Among 5,506 ADRs in 2,765 patients, 104 (1.9%) were classified as serious, though none were fatal. ADRs were reported in a total of 370 different AM preparations. Patients recovered from the majority of ADRs (67.8%), 0.2% recovered with persisting sequelae, 4,8% of ADRs were documented as unresolved and outcomes were unclear in 19.9% of cases. Reported reactions varied by administration route, with site reactions common in local and parenteral use, while oral formulations frequently caused nausea, rashes, and pruritus. Five hepatobiliary cases were investigated for herb-induced liver injury, though this was deemed unlikely. The ADR rate was 1.5 per million maximum daily administration doses (MDADs), with 0.03 serious ADRs per million MDADs.

### Systematic Reviews

Braunwalder et al. conducted a systematic review (SR) of the efficacy and safety of phytotherapy and AM in seasonal allergic rhinitis ^46^. They identified 3 studies, all of which tested Gencydo (*Citrus limonis (lemon)/ juice & Cydonia oblonga (quince)/fruit*) as a nasal spray or subsutaneous injection and reported improvements in nasal and ocular symptoms. One of the included studies was a case series report ^50^, another used laboratory parameters as the primary and clinical parameters as the secondary endpoint ^51^ and the third was the paper discussed above ^33^. The authors interpreted these results as promising regarding the relief of allergy symptoms.

Ploessler and Martin examined the application of AM in chronic pain conditions, identifying seven studies. ^6^ The authors included three RCTs, one of which was discussed above ^36^, the other report on non-pharmacological interventions.^52,53^ Two reports of the same non-randomized trial ^54,55^ with different follow-up times used a multimodal approach and “*did not follow a standardized protocol*”. ^55^ A third non-randomized study was available as a conference abstract only. ^56^ Furthermore, two pre-post analyses were included, none of which had a control group. ^57,58^ The authors concluded that, due to the paucity of the evidence and high heterogeneity of study designs, effects of AM treatments on chronic pain conditions are unclear.

Schwermer et al. ^47^ reviewed the use of AM in pseudocroup in pediatric patients. They included one retrospective study ^59^ and 5 case reports. ^59–63^ Three of those were published in “Der Merkurstab”, the other two were expert opinions copied from AM books ^62,63^. In the retrospective study 103 patients received the AM remedy Bryonia/Spongia comp. (*Apis mellifica D3, Belladonna 3D Bryonia 3D and Spongia 3D*) as well as *Pyrit (3D or 6D) and lavender oil (10%).* Two patients required steroids, 5 received nebulized adrenalin and one needed intubation. This cohort was compared with data from a different group of similar patients. Since both groups had the same otherwise unspecified “complication rate”, the authors concluded that, by employing AM, the use of steroids can be avoided. The expert opinions cannot be categorized as evidence and are thus not discussed further. While the authors mention that clinical trials demonstrating efficacy are missing, they conclude that there is a broad range of AM remedies indicated for pseudocroup. No quality assessment/risk of bias analysis was reported.

The same group also published a systematic review addressing AM for acute gastroenteritis in children. ^48^ The authors included two reviews ^64,65^, two observational studies ^66,67^ and three ‘experience reports’ ^62,63,68^. All the reviews and trials were published in the AM-journal “*Der Merkurstab*”. Recommended treatments were Potentilla erecta ^65^, Gentiana Magen (stomach, translated from German) Globuli velati ^64,66^ and Bolus alba comp. ^67^. The expert opinions from so-called “grey” literature cannot be categorized as evidence and are thus not discussed further. The authors concluded that data for AM in children is insufficient, since no rigorous trials are available. No quality assessment/risk of bias analysis was reported.

### Risk of Bias and Quality Assessment

All of the included RCTs and non-randomized studies are wide open to selective reporting bias and spin (Tables 3 and 4). Five out of 6 primary endpoints were not reached ^34–38^ with one primary endpoint being significant in the per protocol but not intention to treat analysis.^33^ Considering the high number of protocol violations and drop-outs reported, even the one positive result should be interpreted cautiously. Most of the conclusions are misleading, for instance by interpreting significant secondary findings from sub-group or post-hoc analyses as proof of efficacy.

**Table 3a:**
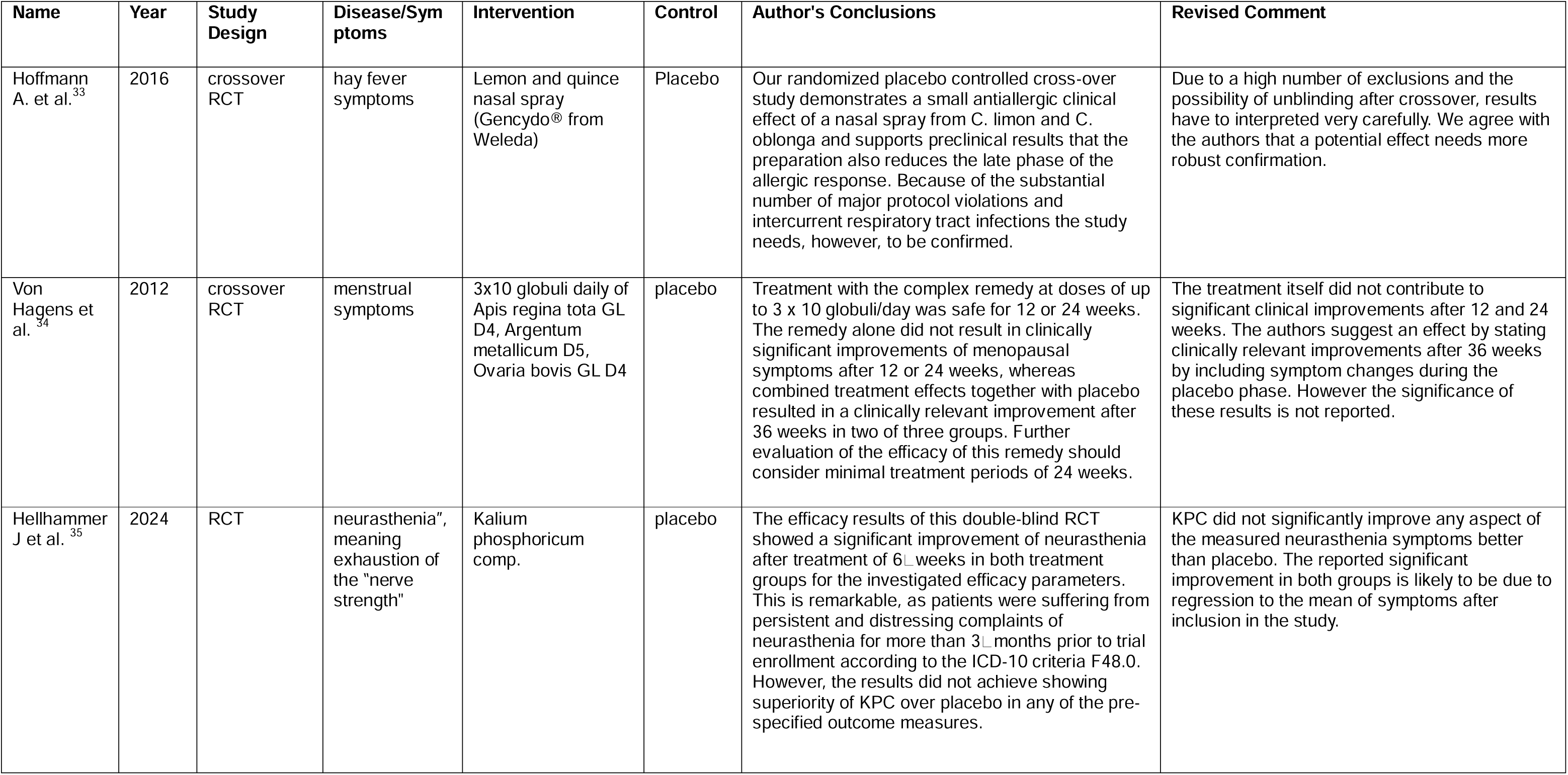
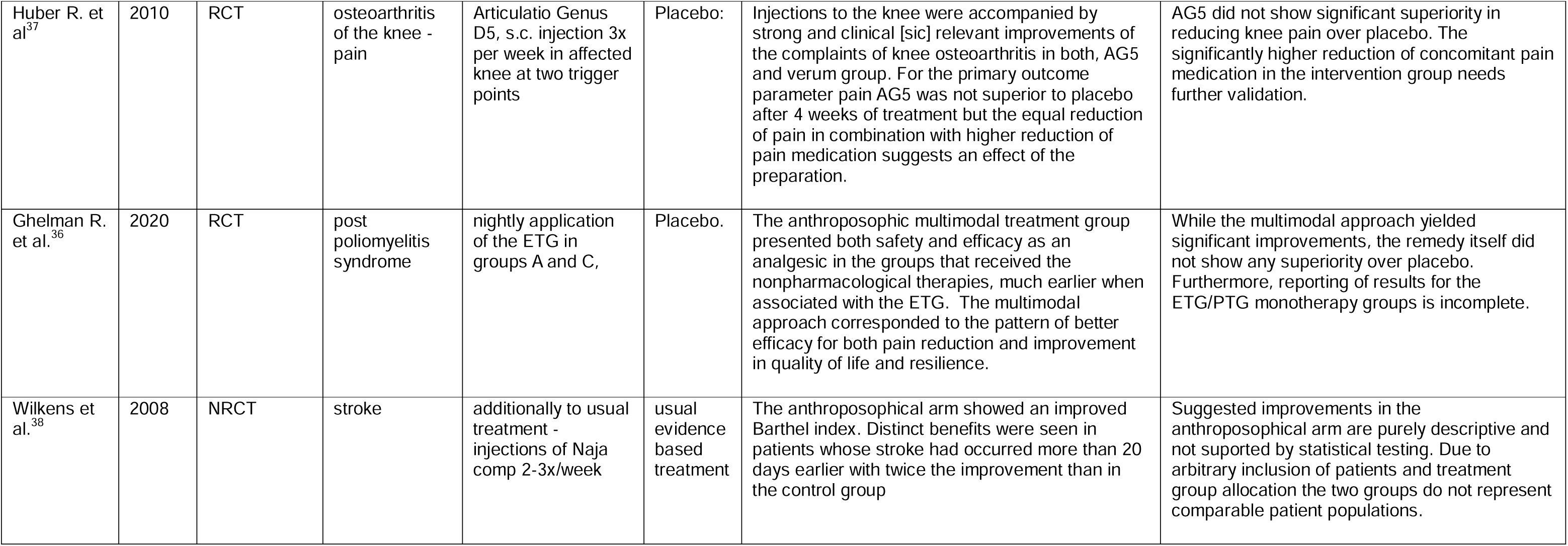
Assessment of presented evidence. Original author’s conclusion vs. our interpretation of intervention studies.

**Table 3b:**
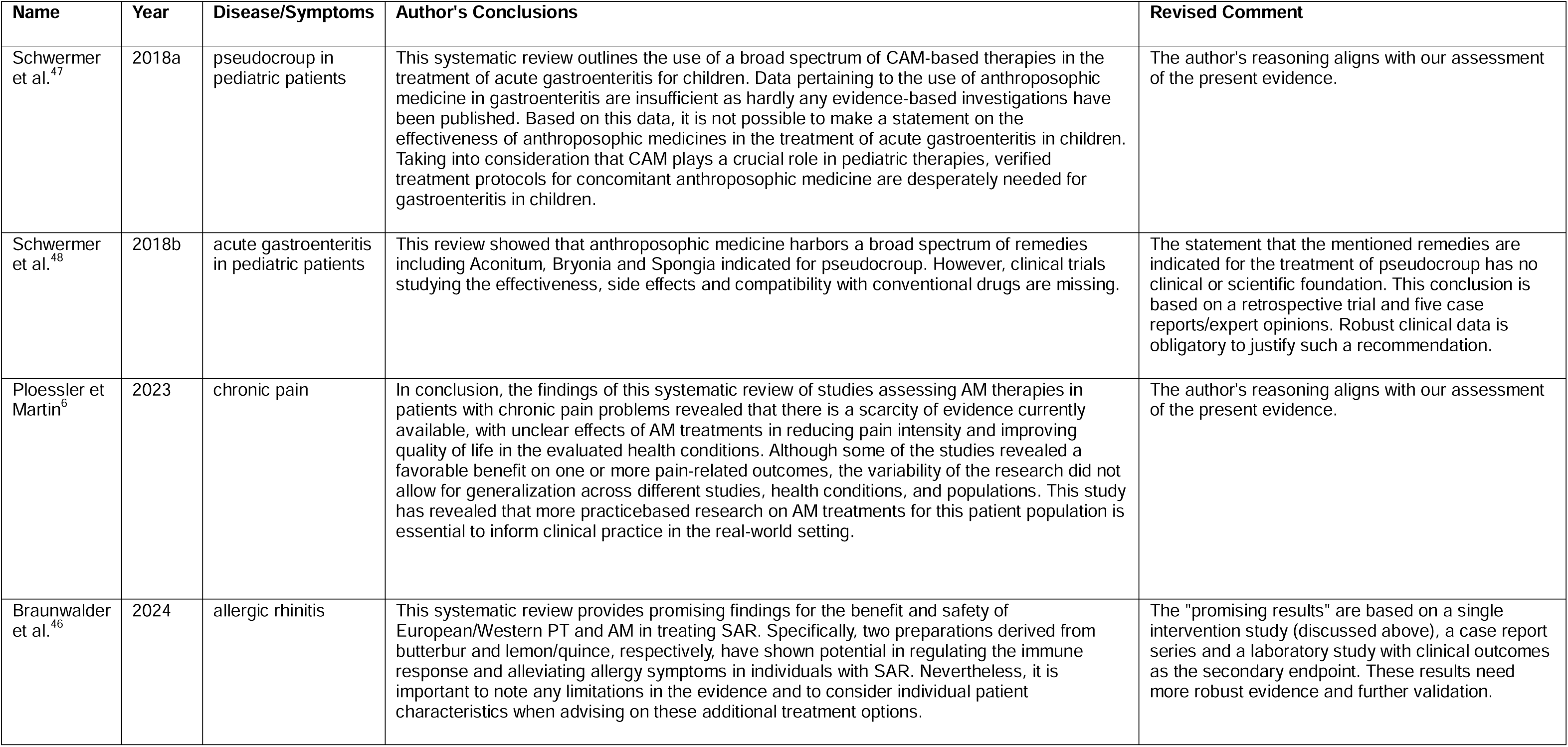
Assessment of presented evidence. Original author’s conclusion vs. our interpretation of systematic reviews.

**Table 3:**
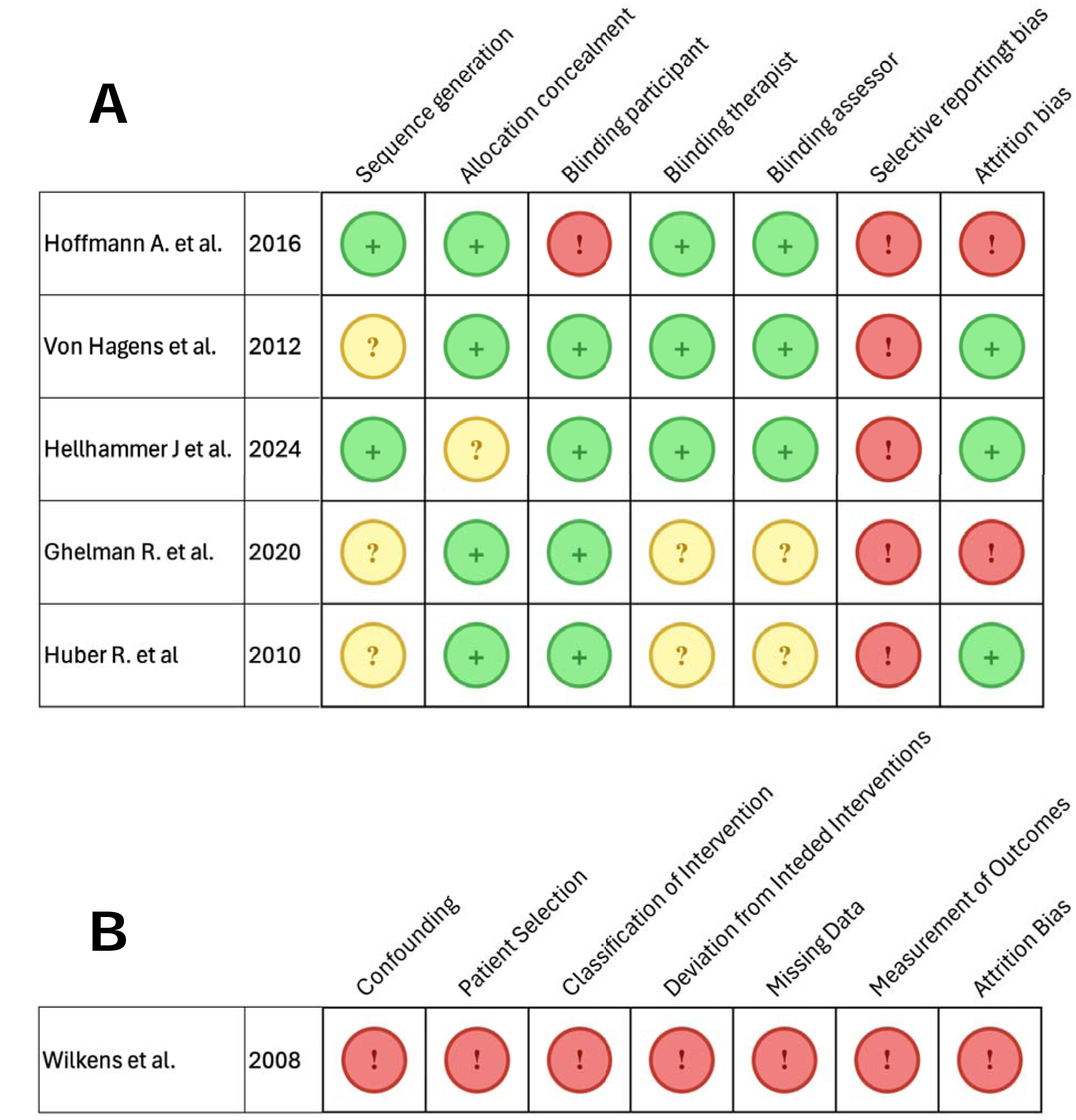
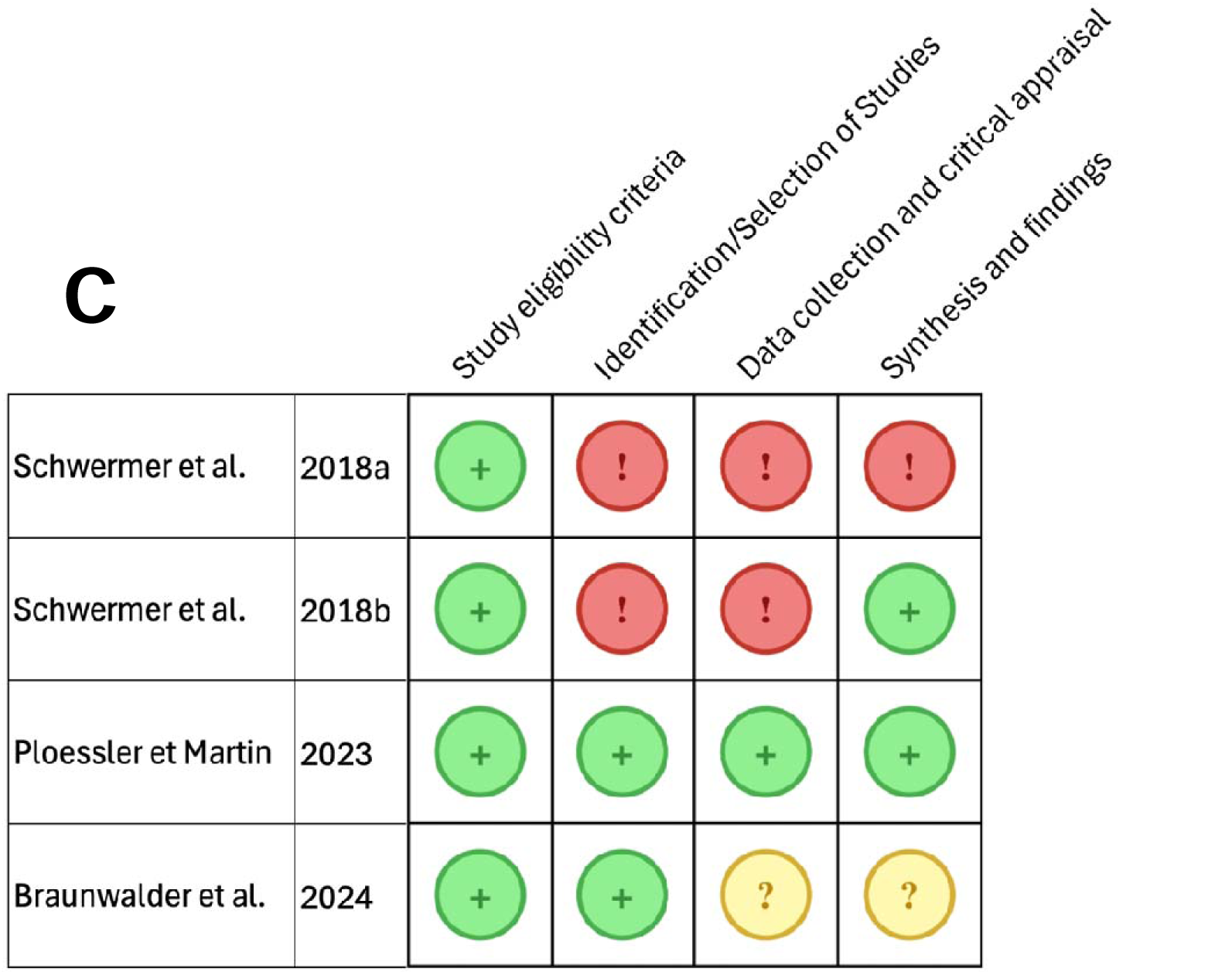
Risk of Bias Assessment of A) randomized trials (RoB-2), B) non-randomized trials (ROBINS-I) and C) systematic re iews (ROBIS)

Allocation concealment and blinding of participants was inconspicuous in most RCTs (Table 4a). One trial had a high risk of participant unblinding due to different tastes of placebo and intervention, i.e. AM administered orally.^33^ Its design implies that after crossover, the difference could have been noticeable.

The risk of inadequate sequence generation^34,36,37^ and unblinded therapists or assessors^36,37^ was unclear in three and two trials respectively. Attrition bias was high in two RCTs^33,36^ due to a high dropout and/or exclusion rate.

One non-randomized study^38^ had an exceptionally high risk of bias. Its patients were selected and allocated by the trialists, the intervention and control group differed greatly in terms of disease severity, baseline scores and results were reported incompletely and there was no statistical testing of the results. None of this stopped the authors of the study to recommend AM for stroke rehabilitation.

Two of the SRs^47,48^ were rated as high risk, due to their inclusion of “grey” literature^2^ and expert opinions. The SR by Braunwalder et al.^46^ was classified as unclear, since its conclusion was based on one trial primarily testing laboratory parameters.

As for prospective ^39–42^ and retrospective reports on ADR’s^45^, the inclusion of participants was open to bias. Often, only doctors practicing AM, were included as researchers. Indeed, Hamre et al. admitted that reporting bias is inherent to their study and noted the necessity of training physicians in ADR-reporting to minimize bias. Similarly, all physicians participating in the study by Tabali et al.^40^ were members of the German Association of Anthroposophical Physicians (GAÄD) and thus not free of pro-AM bias.

## Discussion

Our systematic review aimed to summarize and evaluate evidence for the efficacy and safety of substance-based AM. Only 17 of 360 studies identified could be included, and merely 5 of these were RCTs. None of the RCTs was methodologically sound and none reported unambiguous superiority of AM over control interventions or placebos. Despite the failure to generate evidence in favor of AM, the authors of these RCTs often try to mislead the public by misinterpreting their own findings. Two RCTs initiate their conclusion by highlighting the significant clinical improvement of symptoms *within* both, the intervention and the placebo group. The *between* group comparison/analysis is mentioned secondly, although being the primary endpoint. An example is the following statement: our “*RCT showed a significant improvement of neurasthenia after treatment of 6*LJ*weeks in both treatment groups”*; only later the authors admit that *“the results did not achieve showing superiority* [the AM] *over placebo in any of the pre-specified outcome measures”* ^35^ It is difficult not to get the impression that authors of AM studies tend to be blinded by their own belief in AM and their wish to advertise rather than test AM. These studies excel in demonstrating two well-known effects, that make RCTs imperatively obligatory in clinical practice – self limitation of acute symptoms and regression to the mean of acute and chronic symptoms. Correct interpretation of such results requires a thorough understanding of these concepts and their implications.

### Safe drugs or lack of pharmacological action?

Four prospective and one retrospective study provided data on AM safety. Collectively, they reported an extraordinarily low incidence of ADRs and outstanding patient satisfaction. Considering that AM medications are heavily inspired by the dilution and potentiation processes of homeopathy, these numbers are neither surprising nor unexpected.^69^ As noted by the German pharmacologist Gustav Kuschinsky: “*A medicine that is claimed to have no side effects is under strong suspicion of also having no main effect.*” ^70^ (Original text, German: “*Ein Arzneimittel, von dem behauptet wird, daß es keine Nebenwirkungen habe, steht im dringenden Verdacht, auch keine Hauptwirkung zu besitzen.*”) The non-superiority of homeopathic medications over placebo has been well documented.^71^ Similarly, there is no reason to assume a different outcome by administering these under a different label. Considering the impressively low incidences of reported ADRs prompts the question: What is the baseline incidence of the reported symptoms in the untreated or placebo population. The incidence of ADRs even of an inert placebo is known to be considerable. Around one-quarter of all patients who receive a placebo in clinical trials even stop taking it due to the adverse effects caused by placebos^72^. It is therefore hard to understand why the rate of ARDs should be considerably lower for AM. In a systematic review Rief et al.^73^ analyzed the incidences of, typically unspecific, adverse events in the placebo groups of statin trials and compared them to a sample representing the general German population. They observe an effect were some reasonably prevalent unspecific symptoms (e.g. headache, nausea, fatigue) are underreported in clinical trials, possibly due to a positive drug attitude of volunteers. However, they also report a high variance of ADR incidences between trials. The striking difference compared to our reviewed data is the discrepancy of several tenfold decreases in AM-related ADRs in RCTs as well as in pharmacovigilance data. This leads to the conclusion that either symptoms are underreported in anthroposophical settings, or the causality assessment of nonspecific symptoms is heavily biased against AM.

### Systematic reviews – what is being reviewed?

Four SRs met our inclusion criteria. However, considering the evidently slowly developing field of publications in anthroposophical medicine the question arises: Where is the data coming from? What is even being reviewed? Both reviews by Schwermer et al.^47,48^ included several “expert opinions” which are on the lowest level of evidence hierarchy. The number of “expert opinions”, with five out of six^47^ and three out of seven^48^ of the included papers, respectively, is exorbitantly high. Schwermer et al. even state that there is a broad spectrum of remedies *indicated* for the treatment of pseudocroup, but conclude this statement with a call for clinical trials.^47^ The scarcity of evidence is further demonstrated by the low number of included trials in the other SR as well. Braunwalder et al. ^46^ found only three publications investigating AM in the treatment of allergic rhinitis and only one was a clinical trial. While the authors do not interpret this as evidence for clinical efficacy, they do conclude these results to be promising. Altogether the SRs did therefore not meaningfully contribute to the evidence.

### A comment on study design and statistical analysis

Randomized studies are the accepted standard for testing the efficacy of medical interventions. In view of the criticism voiced against the SR by Ernst 20 years ago,^3^ we nevertheless included non-randomised trials in our review and were more lenient to expand the scope of our search beyond the current gold standard to demonstrate clinical efficacy. To this end, the overwhelming majority of published trials could not be considered in any way, owing to their missing control groups. While we recognize the importance of single arm trials in hypothesis generation, preliminary testing and pharmacovigilance studies, their results cannot be used to draw any causal conclusions. This includes the often-cited AMOS study, which included no control group.^74,75^ By failing to control for (i) the placebo effect and unblinded conditions (two of the most crucial biases), (ii) omitting any randomization, and (iii) ignoring regression to the mean and the self-limitation of symptoms, these studies cannot be ethically interpreted in any clinical setting.

The only non-randomized trial that was included by Wilkens et al. ^38^ administered Naja comp. to patients admitted to a rehabilitation center after suffering a stroke. Only patients from one of two centers were considered for this review, due to the multimodal treatment in patients from the other center. Apart from being not randomized, this study had several further flaws. Treatment allocation was entirely arbitrary, unblinded and according to the discretion of the trialist. The intervention and control groups were not comparable, with higher post-stroke impairment in the intervention group. All the primary endpoint data was presented by descriptive statistics and not supported by any testing. The only statistical models that were calculated were multiple regression models of a subgroup to determine the odds for improvement. The fact that the authors nevertheless conclude that Naja comp. is appropriate for the treatment during stroke rehabilitation supports our suspicion that authors of AM studies might not be objective.

The included RCTs strike with irregularities and questionable interpretations of their results. Hellhammer et al. ^35^ randomized patients to receive KPC to treat stress, exhaustion and fatigue in neurasthenia. Besides defining three separate measures for the primary outcome, a convenient practice enabling selective reporting, pain was assessed on multiple Likert scales and analyzed by t-tests, where non-parametric tests would have been more appropriate. Despite not reaching the primary endpoint, in an exploratory post-hoc analysis of the total of 66 subscales, KPC showed significant improvement in three of those: irritability (p = 0.020), nervousness (p = 0.045) and joy (p = 0.037). Assumingly, due to the explorative nature, no correction for multiple testing was done. Ultimately, the trial’s overall results are framed to suggest a reduction in stress related symptoms, given that the other sub-scores had non-significant but “greater improvements” compared to placebo. They further conclude that using overall improvement as an endpoint, instead of mean values at follow-up would demonstrate the superiority of the drug. However, assuming proper randomization, blinding and analysis such an effect should be reflected in either approach.

Recent decades have seen considerable progress in healthcare such as a general acceptance of the principles of evidence-based medicine. Breakthrough discoveries such as the antibiotic properties of penicillin, the isolation of paclitaxel from the bark of the Pacific yew or the severe birth defects following thalidomide treatment in pregnant women have taught us invaluable lessons about natural and synthetic drugs as well as careful study design and close pharmacovigilance. These lessons prompted the development of our current gold standards of Good Clinical & Scientific Practice, a rigorous drug approval process and the importance of well-designed randomized clinical trials to ensure minimal risk of mistaking correlation for causation when interpreting novel clinical data. Yet some CAM advocates have argued against these achievements and advocated new standards of evidence and research designs specifically for CAM treatments. ^76^ On this background, it seems remarkable that the present review arrives at strikingly similar conclusions as the one published 20 years ago.^3^ This striking lack of progress might indicate that, within the realm of AM, new standards and research designs have not been forthcoming.

### The plausibility of AM

While evidence-based medicine can benefit from rigorous and thoughtful criticism to keep ever improving its metaphysical (i.e. a priori) presuppositions, it appears imprudent not to consider closely why it was established in the first place. Most esoteric metaphysical frameworks of CAM-treatments inherently lack rigorous epistemic virtues altogether – and, most importantly, as a result, lack the immense explanatory power of the currently established paradigm of evidence-based medicine.

In this review we demonstrate how instrumentalization of several biases is applied to tell stories and introduce recommendations in a “medical sub-discipline” that repeatedly fails to withstand the challenges of the simplest clinical trials. In keeping with our explanation in the introduction, the assumptions of AM fly in the face of science. In view of the present evidence and lack of impactful progress over the past 20 years, this use of allocated research funds appears irresponsible. Unless substantial methodological considerations and improvements are implemented, we fear that research funds might be waisted, if AM is submitted to more research of this quality.

### Strengths and limitations of this study

Being aware of the criticism voiced against a previous review,^3^ our SR deliberately set out to be more inclusive. Thus, we did not focus merely on RCTs, conducted more extensive literature searches and included more recent papers. Yet, we can never be entirely sure that we did not miss relevant studies. In particular, we were restricted by our knowledge of just 6 languages, and it seems possible (however, we think unlikely) that more evidence has been published in languages excluded here and not listed in any of the electronic databases we used.

Another criticism that might get voiced is that our team of authors is biased against AM. We gladly admit that we are critical and stress that it is the purpose of an SR to be critical. However, we reject any accusation of anti-AM bias and point to the fact that this review was conducted transparently to minimize bias wherever possible.

## Conclusion

The current SR fails to generate convincing evidence for the efficacy of substance-based AM treatments. Furthermore, it suggests that the ADR rates for AM are even lower than those expected of placebos. This extraordinary finding could be explained by underreporting of ADRs in the anthroposophic setting and an insufficient/unreliable causality assessment; furthermore, there might also be an effect of care (“Placebo by attention”). While it remains to be answered whether the non-pharmacological interventions of anthroposophical medicine aid in the rehabilitation of acute and chronic conditions, we conclude that there is insufficient evidence to support the use of substance bound anthroposophical preparations in any setting.

## Data Availability

Further data is available in the supplementary files. All data produced in the present study are available upon reasonable request to the authors.

## Reporting guidelines

A checklist for reporting systematic reviews needs to be uploaded. It is based on the PRISMA guidelines and sets out the information readers need to fully understand your work.

## Author Contributions

HHS conceived the idea for this review. HHS, MCGP, DS and JH conceptualized the manuscript. DS wrote the initial draft. DS, NR, FT, JO, PT, JZ, HHS prepared and wrote the manuscript. DS, NR, FT, JO, JZ, HHS, AR, JH, EE and MCGP reviewed the manuscript, DS, NR, FT, HHS, JH and EE edited the manuscript. DS, NR, JO and PT reviewed the literature and extracted the data. DS supervised and organized the literature review and data extraction. HHS, MCGP, JH and EE were project supervisors. All authors have read and agreed to the submitted version of the manuscript.

## Author statement

The authors declare no competing interests.

## Data statement

Data extraction tables and other materials are available in the supplementary information. For any other information, please contact the corresponding author.

## Funding

Supported by the Medical University of Vienna, Vienna, Austria.

## Registration

This Study was registered in PROSPERO with the number PROSPERO 2024 CRD42024620083, available from: https://www.crd.york.ac.uk/prospero/display_record.php?ID=CRD42024620083. No amendments to the original protocol were necessary.

## Acknowledgements

We cordially thank Prof. Herwig Czech for critically reading and reviewing the manuscript, especially in with respect to the historical contextualization of our study.

## Footnotes

1 In this context, the RCT refers specifically to a two-arm, randomized, placebo-controlled phase II trial.

2 „grey literature” denotes material and research produced by organizations outside of the traditional commercial or academic publishing and distribution channels.

## Notes

### Competing Interest Statement

The authors have declared no competing interest.

### Clinical Protocols

https://www.crd.york.ac.uk/prospero/display_record.php?ID=CRD42024620083

### Funding Statement

This study was supported by the Medical University of Vienna, Vienna, Austria.

### Author Declarations

This systematic review used only data of already published human trials from established data bases. (Medline, EMBASE, Cochrane Central Register of Controlled Trials, CINAHL, PsycInfo, and Anthromedlit-Datenbank)

