## Supplementary material for "Efficacy and Safety of Substance-Based Therapies in Anthroposophical Medicine: A Systematic Review": Supplements Selection Criteria and Outcomes.docx

|  | **INCLUSION CRITERIA** | **EXCLUSION CRITERIA** |
| --- | --- | --- |
| **POPULATION:** | Adults and children of all ages | Not applicable |
| **INTERVENTION:** | Anthroposophic substance-based treatment. | - Mistletoe treatment - Other treatments: artistic therapies, eurythmic movement exercises, physiotherapy modalities as rhythmical massage therapy and nursing techniques, Integrative therapy approaches with several distinct interventions |
| **CONTROL:** | - Placebo - other active substance | No treatment, no control group |
| **OUTCOMES:** | - Morbidity, related to:   - Cardiovascular outcomes   - Cognitive outcomes   - Behavioural outcomes   - Adverse events (e.g. nausea, allergic reaction, headache, gastrointestinal problems, kidney failure, hospitalization, therapy discontinuation) - Mortality, related to:   - Cardiovascular outcomes   - Adverse events (e.g. nausea, allergic reaction, headache, gastrointestinal problems, kidney failure, hospitalization, therapy discontinuation) - Composite endpoints (related to the criteria mentioned under Morbidity & Mortality - Other patient relevant outcomes (e.g. quality of life, activities of daily living) | - Surrogate outcomes (e.g. laboratory results) |
| **TIMING:** | No cutoff/limitations | No cutoff/limitations |
| **SETTING:** | No limitations (in hospitalized patients/outpatients) | No limitations |
| **STUDY DESIGNS:** | - Systematic Reviews with or without meta-analysis - Randomised clinical trials (RCTs) - Non-randomized controlled studies - Observational studies (prospective and retrospective cohort studies, case-control studies, cross-sectional studies) - Non-controlled pre-post interventional studies | - All other study designs (editorials, narrative reviews, case reports, case series, letters, expert opinions, conference abstracts, qualitative studies, studies with n<30) - duplicates - republished data |
| **LANGUAGES:** | English, German, Spanish, Dutch, Czech, Ukrainian | Other languages |

### Main and additional outcomes

All studies investigating direct patient relevant outcomes are eligible.

- Primary endpoints: Effectiveness of anthroposophic medicine therapy in a particular disease, according to a standardized and validated scale (see above; in brief, including but not limited to: mortality, morbidity, cardiovascular outcomes, cognitive outcomes, and behavioural outcomes.
- Secondary endpoints: adverse events and/or side effects during anthroposophic therapy, other patient relevant outcomes (e.g. quality of life, activities of daily living), as well as composite endpoints mentioned above. Consideration will be given to participants experiencing adverse events of any kind.

Studies investigating surrogate parameters, like laboratory parameters, will be excluded.
